# Public Perception of COVID-19 Vaccines on Twitter in the United States

**DOI:** 10.1101/2021.10.16.21265097

**Authors:** Zidian Xie, Xueting Wang, Yan Jiang, Yuhan Chen, Shengyuan Huang, Haoxuan Ma, Ajay Anand, Dongmei Li

## Abstract

**Background:** COVID-19 vaccines play a vital role in combating the COVID-19 pandemic. Social media provides a rich data source to study public perception of COVID-19 vaccines.

**Objective:** In this study, we aimed to examine public perception and discussion of COVID-19 vaccines on Twitter in the US, as well as geographic and demographic characteristics of Twitter users who discussed about COVID-19 vaccines.

**Methods:** Through Twitter streaming Application Programming Interface (API), COVID-19-related tweets were collected from March 5^th^, 2020 to January 25^th^, 2021 using relevant keywords (such as “corona”, “covid19”, and “covid”). Based on geolocation information provided in tweets and vaccine-related keywords (such as “vaccine” and “vaccination”), we identified COVID-19 vaccine-related tweets from the US. Topic modeling and sentiment analysis were performed to examine public perception and discussion of COVID-19 vaccines. Demographic inference using computer vision algorithm (DeepFace) was performed to infer the demographic characteristics (age, gender and race/ethnicity) of Twitter users who tweeted about COVID-19 vaccines.

**Results:** Our longitudinal analysis showed that the discussion of COVID-19 vaccines on Twitter in the US reached a peak at the end of 2020. Average sentiment score for COVID-19 vaccine-related tweets remained relatively stable during our study period except for two big peaks, the positive peak corresponds to the optimism about the development of COVID-19 vaccines and the negative peak corresponds to worrying about the availability of COVID-19 vaccines. COVID-19 vaccine-related tweets from east coast states showed relatively high sentiment score. Twitter users from east, west and southern states of the US, as well as male users and users in age group 30-49 years, were more likely to discuss about COVID-19 vaccines on Twitter.

**Conclusions:** Public discussion and perception of COVID-19 vaccines on Twitter were influenced by the vaccine development and the pandemic, which varied depending on the geographics and demographics of Twitter users.

## Introduction

The coronavirus disease 2019 pandemic (COVID-19 pandemic) led to serious morbidity and mortality across the US, which is disproportionately affecting racial and ethnic minority populations and elderly people [1] [2]. To achieve herd immunity, it is estimated that at least 70% of the population need to be vaccinated [3] [4]. High acceptance of COVID-19 vaccines will be critical to reach herd immunity [5]. One previous nationally representative survey study showed that the self-reported likelihood of getting a COVID-19 vaccine declined from 74% in early April to 56% in early December 2020 [6]. With the release of related policies and clinical studies of vaccines, the discussion about the COVID-19 vaccine was widespread online [7] [8]. With the development of COVID-19 pandemic and active discussion related to COVID-19 vaccines, it is of importance to monitor public discussion, especially perception of COVID-19 vaccines. While a couple of survey studies have investigated the potential acceptance rate of COVID-19 vaccines [9] [10], the longitudinal and timely surveillance of vaccine acceptance becomes essential during the COVID-19 pandemic.

Social media provides a reliable data source for understanding public opinions toward this COVID-19 pandemic. Social media usage becomes increasingly popular during the COVID-19 pandemic, which provides us a unique opportunity to provide real-time monitoring of public discussion of the COVID-19 pandemic, especially public perception of COVID-19 vaccines. Twitter, as one of popular social media platforms, has been widely used for sharing public opinions on some important public health issues especially the COVID-19 pandemic [11] [12]. Several studies have examined the discussion and public perception (such as vaccine hesitancy) of COVID-19 vaccines using Twitter data [13] [8] [14] [11]. While most of these studies focused on public discussion and perception of COVID-19 vaccines, there is limited data on understanding their potential geographic and demographic differences.

In this study, using Twitter data, we examined the discussion and public perception of COVID-19 vaccines in the US longitudinally. More importantly, by employing computer vision techniques, we estimated basic demographics (including age, gender and race/ethnicity) of Twitter users who tweeted about COVID-19 vaccines, which help us understand public discussion and perception of COVID-19 vaccines among different demographic groups.

## Methods

### Data collection and pre-processing

COVID-19-related tweets between March 5, 2020 and January 25, 2021 were collected through Twitter streaming API using COVID-19-related keywords, including “covid19”, “covid”, “coronavirus”, and “ncov” [15] [16]. Due to technical issues, we failed to collect relevant tweets between August 24, 2020 and September 14, 2020. To extract COVID-19 vaccine-related tweets, we further filtered the COVID-19 tweets using vaccine-related keywords (including “vaccine”, “vaccinated”, “vaccination”, “Pfizer”, “moderna”, “astrazeneca”, “janssen”, “novavax”, and “mrna”). Then, the duplicate tweets and commercial tweets were further removed as described before [17] [18], which resulted in 18,117,984 tweets related to COVID-19 vaccines. Based on the geolocation information provided in each tweet [19], we identified 4,438,203 COVID-19 vaccine-related tweets posted by 1,153,299 Twitter users from the US (Supplemental Figure 1).

### Longitudinal and geographic analysis of tweets related to COVID-19 vaccines

To monitor the discussion of COVID-19 vaccines on Twitter over time, the total number of COVID-19 vaccine-related tweets each week from the US during the study period was plotted. To understand if there is any geographic difference in Twitter users who discussed about COVID-19 vaccines, we calculated the total number of distinct Twitter users in each state and normalized to the state population. In addition, we calculated the average number of COVID-19 vaccine-related tweets posted by Twitter users in each state.

### Sentiment analysis

To understand public perception of COVID-19 vaccines on Twitter, we performed sentiment analysis using VADER algorithm (Valence Aware Dictionary and sentiment Reasoner) [20]. This algorithm uses a dictionary to give a certain text a sentiment score, which could represent the text’s attitude over a certain topic. We applied the vaderSentiment package in Python to calculate the sentiment score for each tweet, and calculate average sentiment scores for each week. The sentiment score is between -1.0 and 1.0. High value of positive sentiment score means more positive sentiment while more negative value indicates more negative sentiment. We defined the sentiment score between 0.05 and 1.0 as positive sentiment, -1.0 and -0.05 as negative sentiment, and the rest as neutral sentiment. To examine the difference in public perception of COVID-19 vaccines among US states, we calculated average sentiment score of COVID-19 vaccine-related tweets from each US state.

### Topic modeling analysis

LDA (Latent Dirichlet Allocation) is a statistical model used to mine the text dataset with some topics constituted by a few frequently occurring words or phrases [21]. For the entire dataset, LDA will evaluate the issues and provide the weight of words that belong to those topics. First, we cleaned the noises before converting the text to a readable model. For example, we got rid of information like emails, extra spaces, URLs, and distracting punctuation in the text. Second, we created the bigram and trigram models. The bigram and trigram models are two or three words that often occurred together in the text. Third, we removed the stop words and then lemmatized the terms into a standard form. Finally, we created the dictionary and corpus as LDA input and built the LDA model. The optical number of topics was determined by the highest topic coherence score.

### Demographic inference

To estimate the demographic characteristics of Twitter users, we performed demographic inference using DeepFace algorithm. DeepFace is an algorithm that can recognize and differentiate human figures and classify them by different thresholds [22]. We built the DeepFace model to estimate three demographic features, including race/ethnicity (Non-Hispanic White, Non-Hispanic Black, Non-Hispanic Asian, Hispanic, and others), gender (male and female), and age groups (Age 18-24, Age 25-29, Age 30-49, Age 50-64, and Age 65+). We compared the sentiments towards COVID-19 vaccines among Twitter users with different demographics.

## Results

### Discussion of COVID-19 vaccines on Twitter in the US

To examine the discussion about COVID-19 vaccines on Twitter over time in the US, we calculated the total number of tweets related to COVID-19 vaccines each week between March 5, 2020, and January 31, 2021. As shown in Figure 1, while the discussion of COVID-19 vaccines fluctuated and remained active at the beginning, there were two major peaks at the end of 2020. The first peak showed up around November 9, 2020 when Pfizer announced that its COVID-19 vaccine could be effective up to 90%. The other peak appeared around December 8, 2020 when the number of COVID-19 cases reached fifteen million in the US.

**Figure 1.**
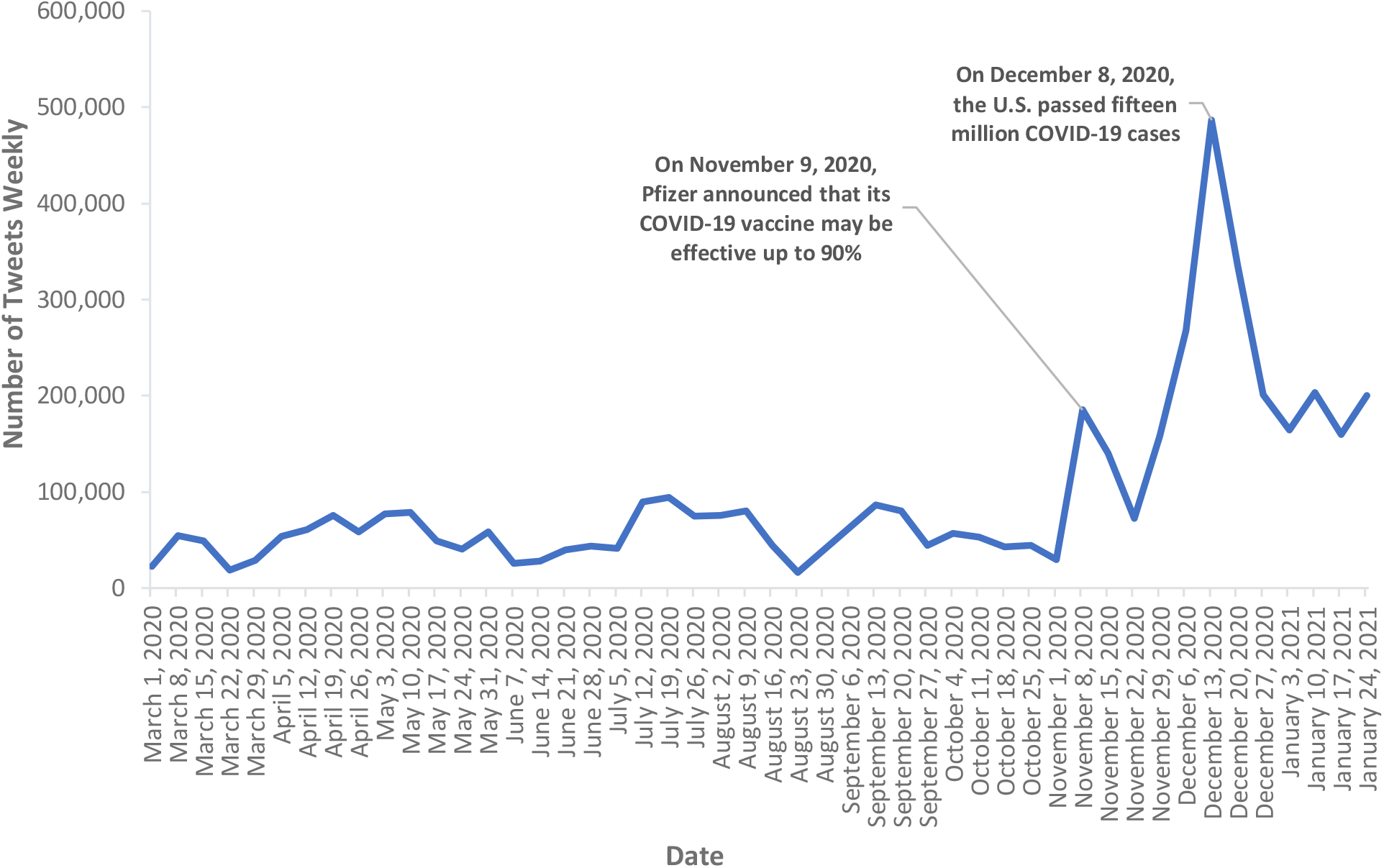
Total number of tweets mentioning COVID-19 vaccines each week in the US.

### Geographic distribution of Twitter users who discussed COVID-19 vaccines in the US

While the discussion of COVID-19 vaccines was active on Twitter in the US, it is important to examine whether there was some geographic difference in discussion of COVID-19 vaccines on Twitter. To address this, we calculated the total number of distinct Twitter users who mentioned COVID-19 vaccines in each state, which has been normalized to the state population. As shown in Figure 2, the states with active discussion of COVID-19 vaccines mainly located at the west, east and south areas of the US. For example, California has 391.3 Twitter users per 100,000 people who mentioned COVID-19 vaccines, followed by Washington (370.3 Twitter users/100,000 people) and Texas (362.4 Twitter users/100,000 people). In addition, we calculated average number of COVID-19 vaccine-related tweets per Twitter user in different US states, and we did not observe obvious difference between US states (Supplemental Figure 2).

**Figure 2.**
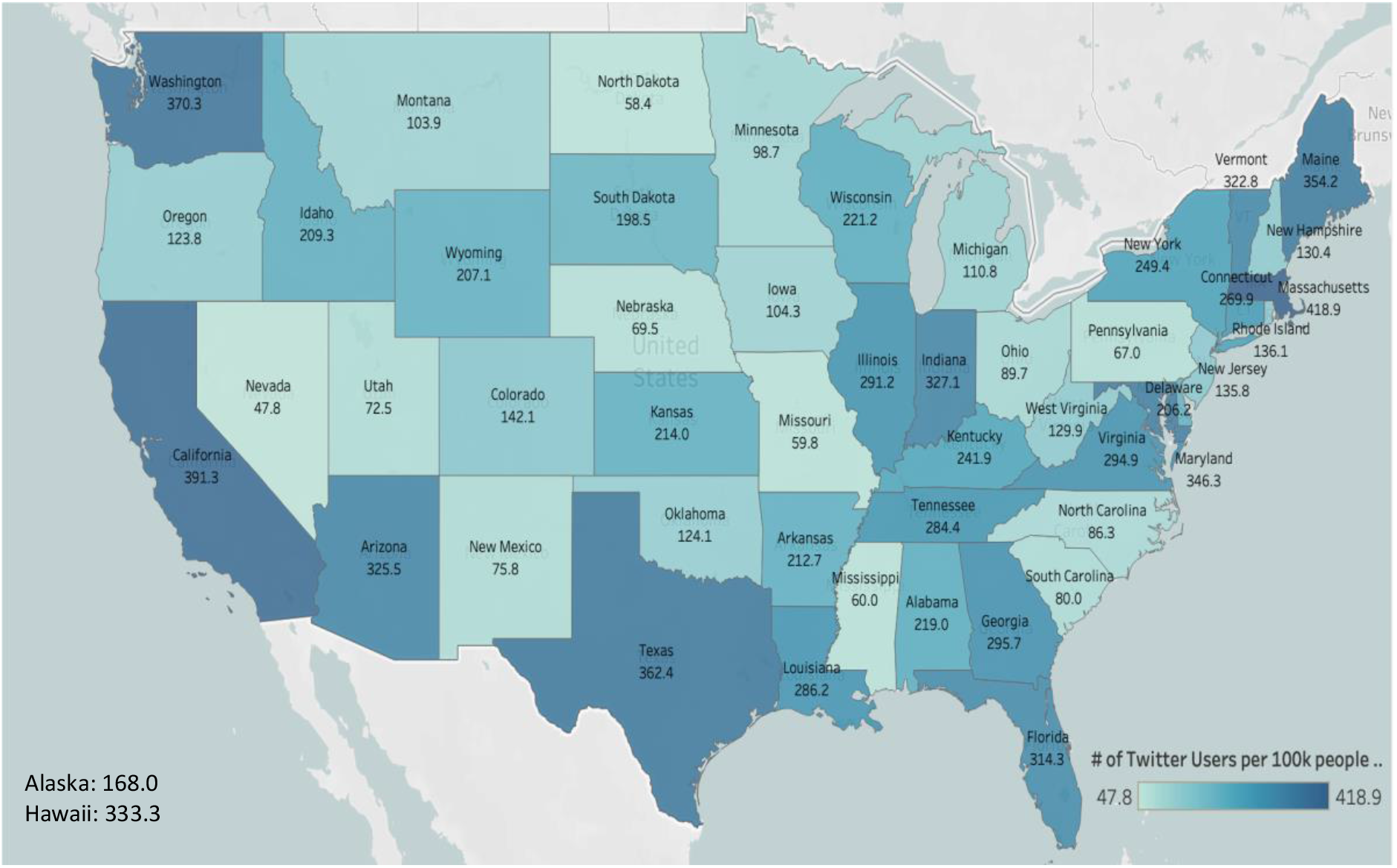
Geographic distribution of Twitter users who discussed COVID-19 vaccines in the US.

### Public perception of COVID-19 vaccines on Twitter in the US

Among 4,438,203 tweets mentioning COVID-19 vaccines in the US, 34.78% (1,543,397) of them had positive sentiment, 32.40% (1,438,107) had negative sentiment, and 32.82% (1,456,699) had neutral sentiment. To understand how the public perceived the COVID-19 vaccines over time, we examined the average sentiment score of tweets mentioning COVID-19 vaccines longitudinally. As shown in Figure 3, average sentiment scores each week varied significantly at the beginning of the COVID-19 pandemic, and became relatively stable later on. There were two big peaks at the beginning of the pandemic. The COVID-19 vaccine-related tweets from March 8, 2020 to March 14, 2020 had the highest average sentiment score (0.155), and the number of tweets from the US during this period was 54,364. The tweets from May 31, 2020 to June 6, 2020 had the lowest average sentiment score of -0.351, and the total number of tweets from US in this period was 58,903.

**Figure 3.**
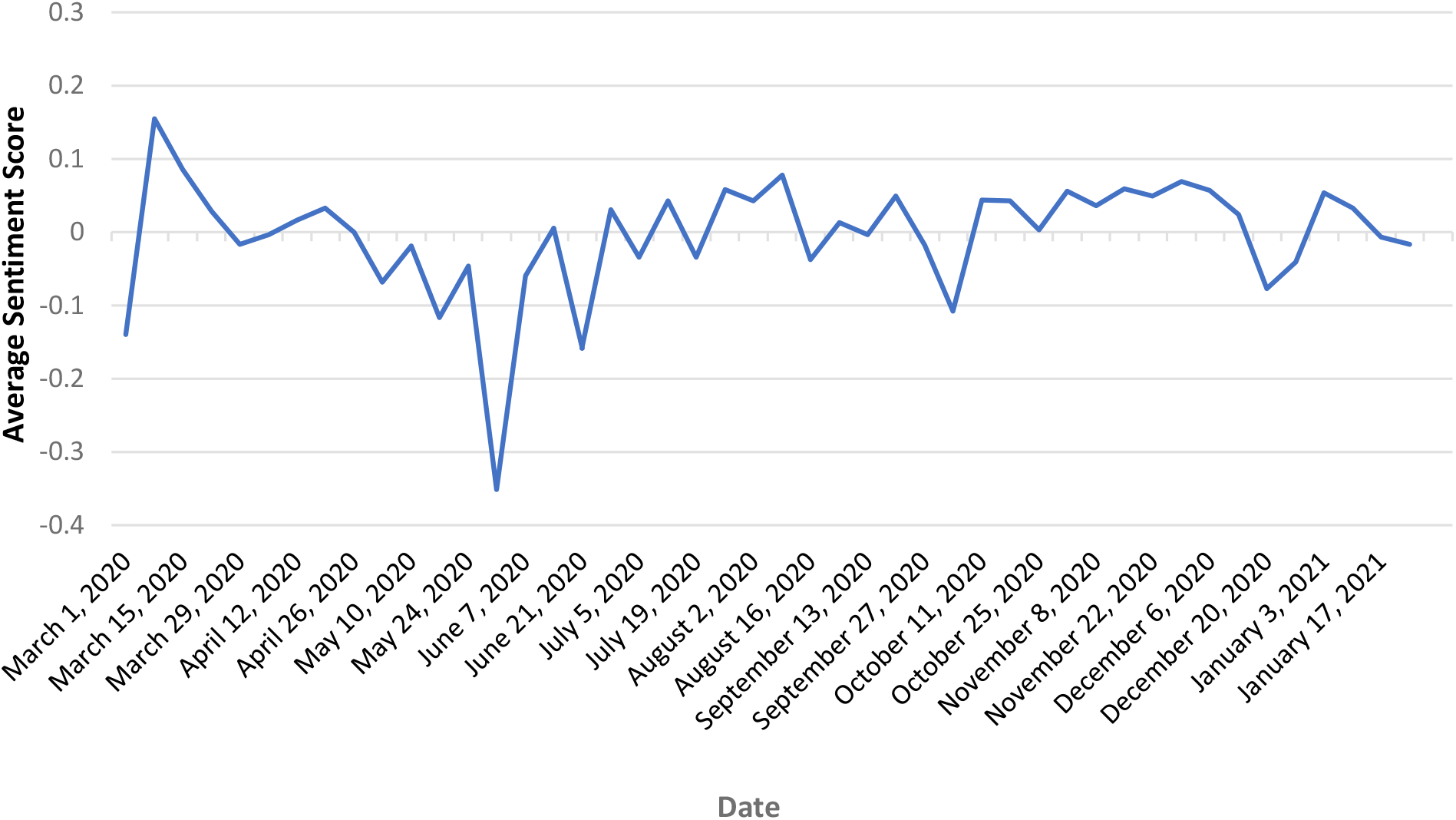
Average sentiment score of tweets mentioning COVID-19 vaccines each week.

In order to understand the potential reasons that lead to these two big peaks in the sentiment score, we performed the LDA topic modeling analysis. Between March 8, 2020, and March 14, 2020, the major topics included “Development of COVID-19 vaccine” (50.43%), “COVID-19 vaccine for everyone” (30.35%) and “Vaccine fight COVID-19 virus” (19.22%) (Supplemental Table 1). Between May 31, 2020, and June 6, 2020, three major topics related to COVID-19 vaccines included “Black lives matter and COVID-19 vaccine” (50.53%), “Bad COVID-19 pandemic” (25.77%) and “COVID-19 vaccine availability” (23.70%) (Supplemental Table 2).

Considering the COVID-19 pandemic varied between different US states, it will be important to examine whether there is some geographic difference in public perception towards COVID-19 vaccines. As shown in Figure 4, average sentiment scores of tweets mentioning COVID-19 vaccines in different states were different. Some states in the east coast had relatively high sentiment scores, for example, Maine (0.025) and New York (0.026). In contrast, some states in the west and mid-west had relatively low sentiment scores, for example, Oregon (−0.014), Colorado (−0.017) and Wyoming (−0.016).

**Figure 4.**
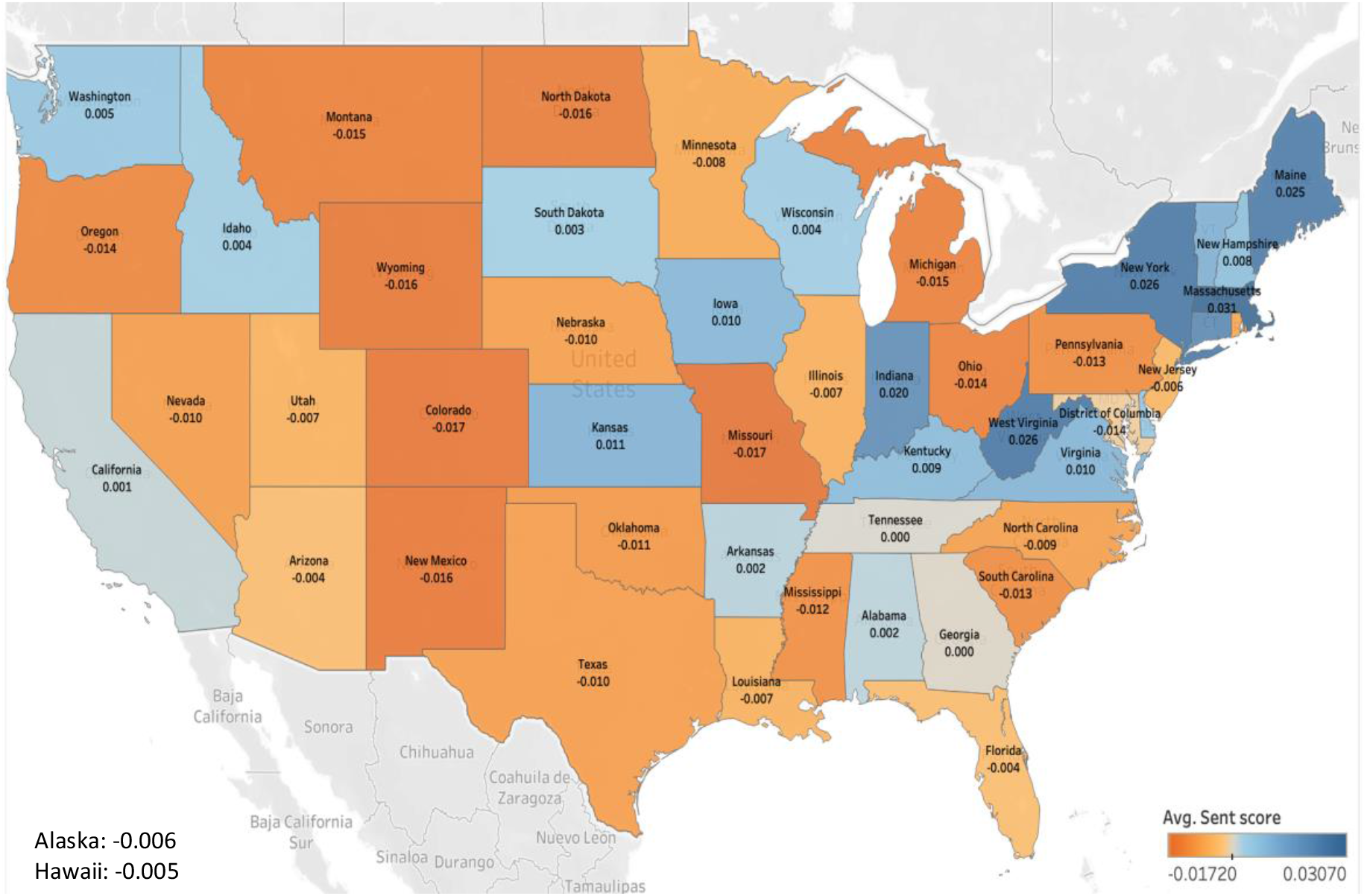
Average sentiment score of tweets related to COVID-19 vaccines from different US states. Avg. Sent score stands for average sentiment score.

### Demographic characteristics of Twitter users who tweeted about COVID-19 vaccines in the US

With an active discussion about COVID-19 vaccines on Twitter, it is of importance to understand who were more concerned about COVID-19 vaccines. We estimated the age, gender, and race/ethnicity of Twitter users who tweeted about COVID-19 vaccines using DeepFace algorithm. The number of unique Twitter users in the US who tweeted about COVID-19 vaccine was 1,135,299 during our study period, among them we successfully extracted 58,823 valid user profile images (5.18%). As shown in Figure 5, there were 38,408 male Twitter users (65.29%) and 20, 415 female Twitter users (34.71%). Among these Twitter users, 30-49 age group was the largest group (66.72%), followed by 25-29 age group (24.01%) and 18-24 age group (6.29%). In addition, among the race/ethnicity groups, White was the largest population (65.18%), followed by Others (16.94%), Asian (8.99%) and Black (5.73%).

**Figure 5.**
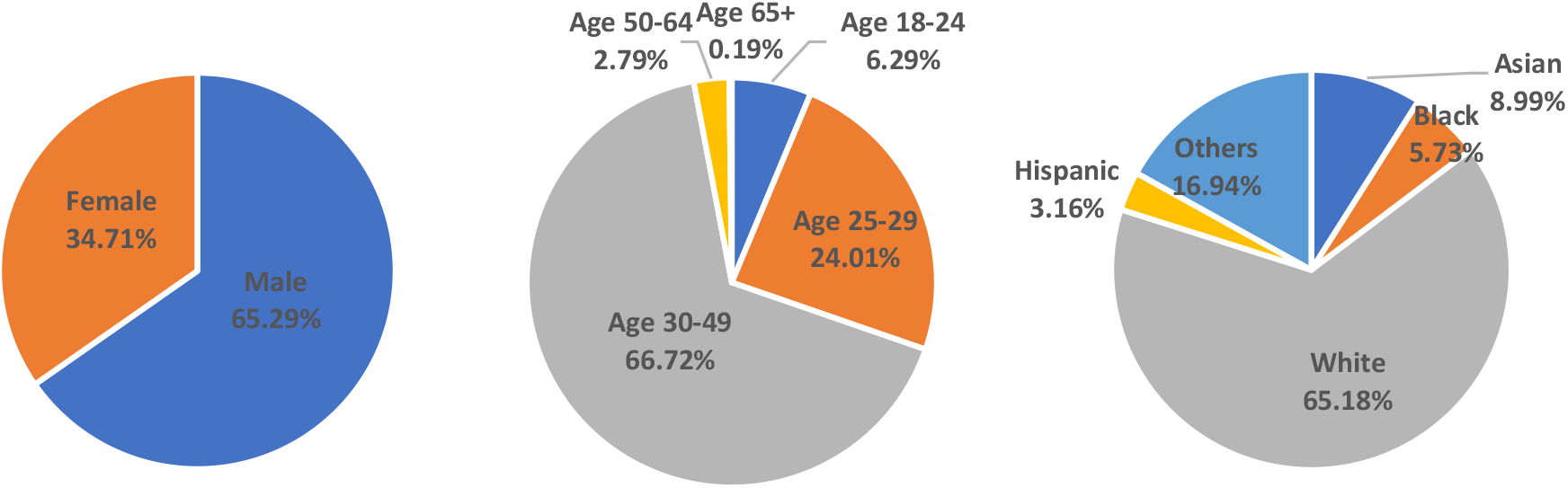
Demographic characteristics of Twitter users who discussed COVID-19 vaccines in the US.

Among these Twitter users, there were 25,962 Twitter users (44.14%) who had positive sentiment towards COVID-19 vaccines, 19,482 Twitter users (33.12%) with negative sentiment and 19,482 Twitter users (22.74%) with neutral sentiment towards COVID-19 vaccines. In addition, we examined whether Twitter users with different demographics had different sentiments towards COVID-19 vaccines. As shown in Supplemental Figure 3, we did not observe any obvious difference in sentiments between different age, gender and race/ethnicity groups.

## Discussion

In this study, we showed the discussion of COVID-19 vaccines on Twitter in the US was increasing over time with some variations, which had been profoundly affected by the COVID-19 pandemic in the US and the development of COVID-19 vaccines. Twitter users from coastal states were more concerned about COVID-19 vaccines. Furthermore, we showed that the perception of COVID-19 vaccines on Twitter in the US varied longitudinally and geographically, and Twitter users from the states in the east coast were more positive towards COVID-19 vaccines. Our demographic analysis showed that the males and 30-49 age group were more likely to mention or discuss the COVID-19 vaccines on Twitter in the US.

In this study, we showed while the discussion about COVID-19 vaccines on Twitter remains very active during the COVID-19 pandemic, there were two significant peaks towards the end of 2020, which is similar to the finding from another study [13]. These two peaks occurred when Pfizer announced that its COVID-19 vaccine could be effective up to 90% [23] and the number of COVID-19 cases reached fifteen million in the US [24], which suggests that the development of COVID-19 vaccines and the severity of COVID-19 pandemic could significantly influence the discussion of COVID-19 vaccines on Twitter.

By examining average sentiment score of COVID-19 vaccine-related tweets over time, we showed that average sentiment score fluctuated around neutral during our study period. In contrast, one previous study showed that the weekly sentiment of COVID-19 vaccine-related tweets remained positive, and increased significantly over time [25]. The discrepancy might result from the fact that different datasets have been used in two different studies, such as our study only focused on tweets from the US, and keywords used for filtering in two studies were different. In this study, we showed that there were more positive tweets than negative tweets, which is consistent with the findings from one previous report [13]. In addition, we observed that there were two big peaks that occurred in early March (positive peak) and late May (negative peak) in 2020. In the week of March 8 to March 14 in 2020, our LDA results showed that there was many positive news about the COVID-19 vaccines, which might drive the positive discussion about COVID-19 vaccines. During the week of May 30 to June 6 in 2020, the top popular topics included BlackLivesMatter and worrying about the availability of COVID-19 vaccines, which might explain the overall negative sentiment of tweets during this time period.

In this study, we estimated basic demographics of US Twitter users who tweeted about COVID-19 vaccines using computer vision technique. Our results showed that the proportion of male in these Twitter users was 65.29%, which is higher than the proportion of male (53.19%) among general Twitter users in the US [26]. Therefore, the male Twitter users in the US might be more likely to discuss or more concerned about COVID-19 vaccines. While 28.72% of general US Twitter users are aged between 30 and 49 [26], 66.72% of Twitter users in our study were aged between 30 and 49, suggesting that Twitter users in 30-49 age group were more likely to discuss COVID-19 vaccines during the COVID-19 pandemic. Compared to general Twitter users in the US [27], the proportion of White in our study was slightly lower (65.18% vs. 68.00%). In contrast, the proportion of Black and Asian in our study was lower than general Twitter users, 5.73% vs. 14.00% and 8.99% vs. 18.00% respectively. These results suggest that the Black and Asian might be less likely to discuss about COVID-19 vaccines on Twitter. By understanding which demographic groups of Twitter users have more concerned about COVID-19 vaccines, we could prioritize specific demographic groups and provide targeted support to ensure their confidence in the COVID-19 vaccines.

There are some limitations in our study. Firstly, even though social media provides a rich data source, it does not provide the demographics of Twitter users. In this study, while we tried to estimate demographics (including age, gender, and race/ethnicity) of Twitter users, there might be some biases due to the accuracy of algorithm. Secondly, considering Twitter users only take up 20% of the US population, the unavailability of posts from private accounts, and many Twitter users did not provide valid geolocation information and profile pictures, our data cannot fully represent the general population. Furthermore, our study only focused on a specific time period during the COVID-19 pandemic, which need to be updated in the future.

## Conclusions

In this study, by mining Twitter data, we examined the public discussion and perception of COVID-19 vaccines spatially and temporally in the US. In addition, we showed that there are some differences in geographics and demographics of Twitter users who tweeted about COVID-19 vaccines. Together, our results provide a comprehensive picture about how COVID-19 vaccines have been discussed and perceived on Twitter in the US.

## Data Availability

All data produced in the present study are available upon reasonable request to the authors

## Acknowledgements

This study was supported by the University of Rochester CTSA award number UL1 TR002001 from the National Center for Advancing Translational Sciences of the National Institutes of Health. The content is solely the responsibility of the authors and does not necessarily represent the official views of the National Institutes of Health.

---

**Supplemental Table 1.** Top topics in COVID-19 vaccine-related tweets from the US between March 8, 2020 and March 14, 2020.

| Topic | Keywords | Percentage of Topics | Examples |
| --- | --- | --- | --- |
| Development of COVID-19 vaccine | vaccine, develop, free, ever, testing, scientist, ready, phase, fast, people | 50.43% | Dr. Anthony Fauci: reaching phase 1 of coronavirus vaccine faster than “anyone has ever done literally in the history of vaccinology”. |
| COVID-19 vaccine for everyone | vaccine, say, virus, flu, shoot, get, make, go, corona, people | 30.35% | "Well, that is a crock" Bernie says about Trump not guaranteeing Corona vaccine will be affordable. "We have got to make sure that when that vaccine comes out, it is available to everybody in this country, regardless of their income." #CoronavirusPandemic |
| Vaccine fight COVID-19 virus | vaccine, find, help, fight, donate, salary, quarterly, flu, can, test | 19.22% | @realdonaldTrump Donates His Quarterly Salary to Help Fight Coronavirus, Find Vaccine. |

**Supplemental Table 2.** Top topics in COVID-19 vaccine-related tweets from the US between May 30, 2020 and June 6, 2020.

| Topic | Keywords | Percentage of Topics | Examples |
| --- | --- | --- | --- |
| Black lives matter and COVID-19 vaccine | people, black, virus, live, dangerous, matter, none, airborne, start, say | 50.53% | Curfew for blacklivesmatter but none for coronavirus.....black people can't be more dangerous than an airborne virus with no vaccine |
| Bad COVID-19 pandemic | need, say, bad, kill, make, develop, covid, apparently, nationwide, solve | 25.77% | Apparently we didn't need a vaccine to solve coronavirus, we just needed some nationwide riots that got ratings for CNN/MSNBC and POOF the virus went and magically disappeared from being a concern anymore |
| COVID-19 vaccine availability | find, doctor, continue, wait, futility, eventually, vaccine, covid, go, get | 23.70% | “Eventually, doctors will find a coronavirus vaccine, but black people will continue to wait, despite the futility of hope, for a cure for racism,” writes Roxane Gay |

**Supplemental Figure 1.**
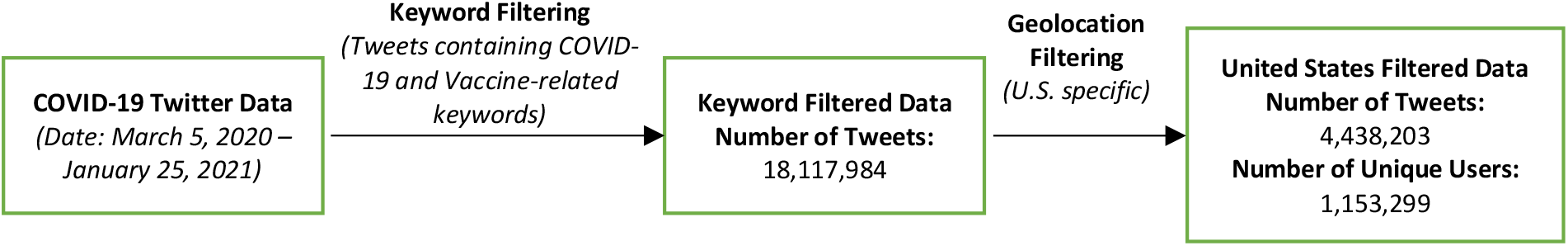
Flow chart of Twitter data pre-processing

**Supplemental Figure 2.**
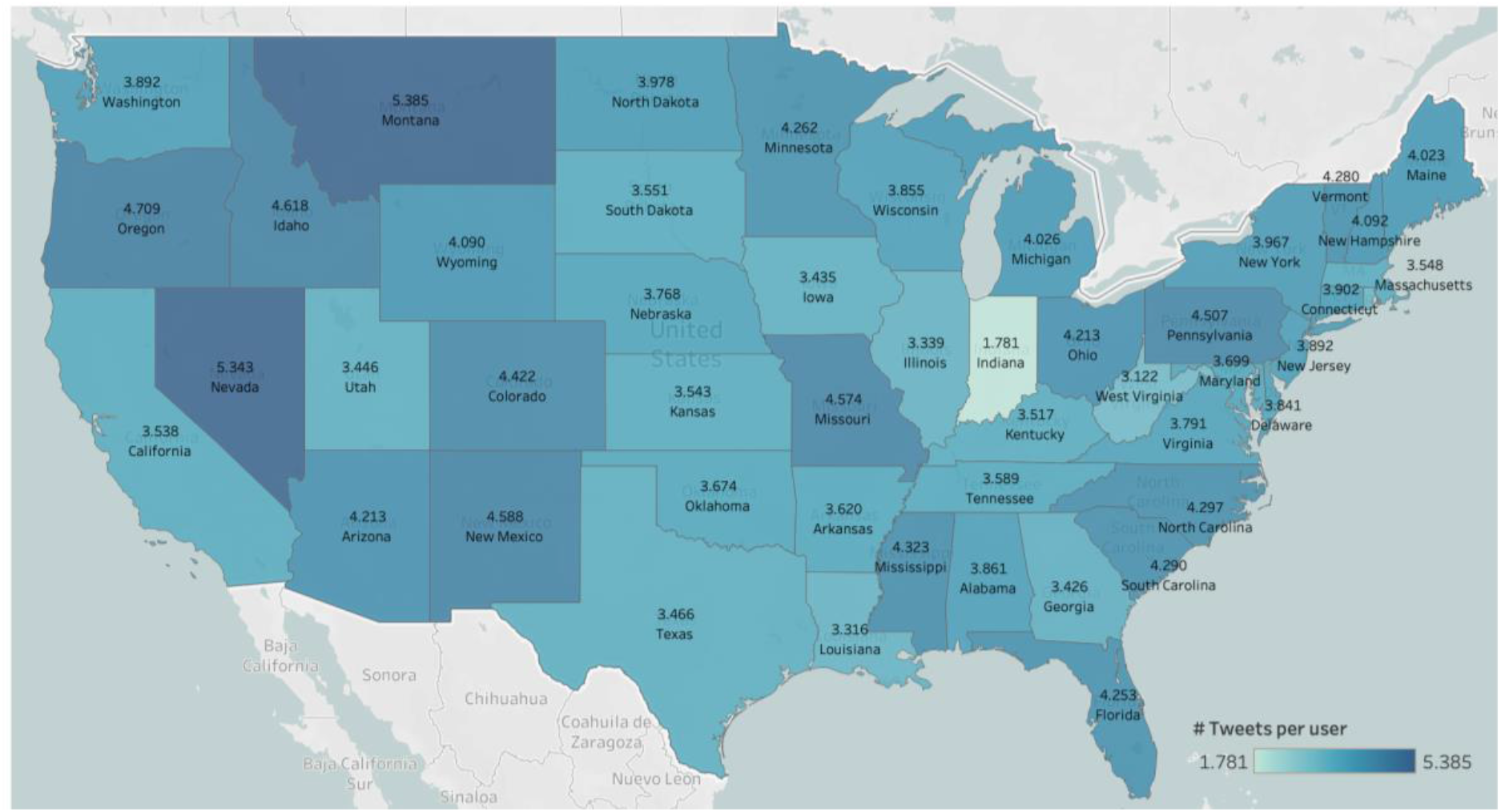
Average number of COVID-19 vaccine-related tweets per Twitter user in different US states.

**Supplemental Figure 3.**
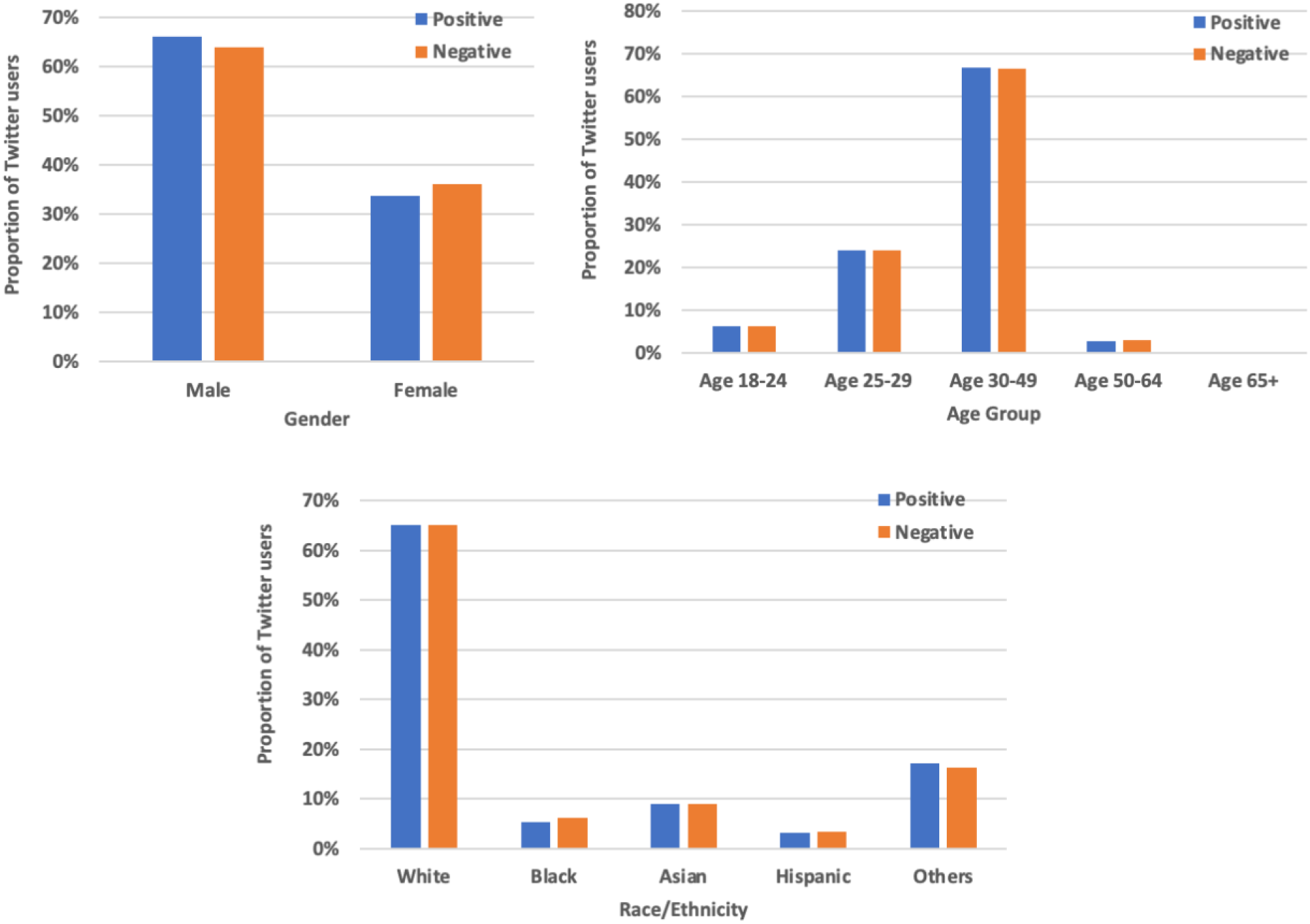
Demographic characteristics of Twitter users with different sentiments towards COVID-19 vaccines.

